# A study of knowledge, experience and beliefs about hepatitis B virus (HBV) infection in south western Uganda

**DOI:** 10.1101/19003129

**Authors:** Joseph Mugisha, Jolynne Mokaya, Dominic Bukenya, Fatuma Ssembajja, Denis Mayambala, Robert Newton, Philippa C Matthews, Janet Seeley

## Abstract

**Introduction:** United Nations sustainable development goals aim for the elimination of viral hepatitis as a public health threat by 2030, leading to efforts to upscale the availability and accessibility of hepatitis B virus (HBV) vaccination, diagnosis and treatment globally. However, a variety of societal factors, including beliefs, traditions, and stigma, can be a major obstacle to all of these interventions. We therefore set out to investigate how HBV is understood and described in communities in Uganda, and whether there is evidence of potential stigma.

**Method:** We carried out a qualitative formative study in two sites in South Western Uganda: a village in Kalungu district (site A) and an area on the outskirts of Masaka town (site B). We undertook a rapid assessment to investigate how adults describe HBV infection and their perceptions about the infection. We collected data by conducting a transect walk, observations, community group discussions, and in-depth interviews, sampling a total of 131 individuals. We used inductive content analysis to extract key themes associated with HBV.

**Results:** There is no specific word for HBV infection in local languages, and knowledge about this infection is varied. While some individuals were completely unfamiliar with HBV infection, some had heard of HBV. Radio was a common source of information. There was awareness of HBV as a cause of liver disease, limited knowledge regarding the cause, mode of transmission and treatment. Stigma in HBV may be rare in this community due to limited understanding and experience of HBV.

**Conclusion:** There is an ongoing need to improve awareness and understanding of HBV in this community. Careful dissemination of accurate information is required to promote acceptance of interventions for prevention, diagnosis and treatment.

## INTRODUCTION

Globally, approximately 290 million people are chronically infected with hepatitis B virus (HBV), and more than 800,000 people consequently die each year from liver cirrhosis or liver cancer (1). In response to the United Nations Sustainable Development Goals, which aim to eliminate viral hepatitis as a public health threat by the year 2030 (2), HBV infection is receiving increasing recognition with efforts to upscale prevention, diagnosis and treatment (1,3). However, there are complex barriers to the elimination of this infection in the African subcontinent, including a high prevalence of chronic infection (≥8% in many settings), >90% of those infected being undiagnosed, and poor awareness (4,5).

Stigma and discrimination are recognised in the setting of chronic viral hepatitis infections, but there is very limited research to determine the nature and impact of stigma in communities in Africa (4). To date, only three studies have been carried out in Africa exploring the social impact of HBV. In Zambia, stigma has been reported to be a barrier to HBV disclosure and referral of contacts for testing (6), whereas in Ghana, low levels of knowledge and pervasive misconceptions about HBV contribute towards psychological and social problems among individuals with chronic infection (7,8). There has been no previous work on stigma in HBV in Uganda, although HBV is highly endemic in some regions (prevalence estimates vary from 6-52% (9–11)).

Given the high prevalence of HBV in Uganda, and the emerging evidence of stigma in HBV, we sought to investigate how adolescents and adults in rural and peri-urban communities in Uganda describe HBV infection, their perceptions, beliefs and experiences about the infection and whether there is evidence of stigma.

## METHODS

### Study setting

We conducted the study at two sites in South West Uganda:

i. Site A: This is a village location within the General Population Cohort (GPC). The GPC was established in a rural population in south-western Uganda in Kyamulibwa sub-county of Kalungu district in 1989, with the initial aim of studying the epidemiology of HIV/AIDS in a rural population, but subsequently also being a source of recruitment for other research studies (12,13). The GPC comprises 25 study villages with a total population of approximately 25,000 people. Most of the inhabitants practice small-scale farming. Local tribes are Baganda, Banyarwanda, Barundi, and Banyakole. A number of infectious diseases are prevalent, including HIV (prevalence ∼10%), malaria, hepatitis B and C (HBV prevalence 3% HCV <1%), sexually transmitted infections and respiratory tract infections. Within this sub-county, there are a number of public and private health care facilities and traditional healers. We selected Site A for this study because a previous survey on HBV reported a higher prevalence within this village compared to elsewhere in the GPC (unpublished data).
ii. Site B: This is a peri-urban site in Masaka Municipality, located along the highway that connects Uganda to the Democratic Republic of Congo, Tanzania and Rwanda, with a high concentration of low-cost housing. Most of the inhabitants earn their living from small businesses including motorcycle transport, selling food and running bars; crop farming is also undertaken locally. The tribes represented in this region are Baganda, Banyarwanda, Batoro and Basoga. The HIV prevalence in this setting may be slightly higher than within the GPC, but accurate data are not available. The prevalence of viral hepatitis infection is also not known but is likely to be higher than the GPC, given the concentration of settlements in the area and many inhabitants from different areas of Uganda. There are several private and public health care facilities in this site.

### Study design

Between July and October 2018, we conducted a rapid qualitative assessment to investigate how HBV is described, the experiences and beliefs associated with HBV and the nature of any associated stigma. These rapid methods included transect walks, observations, introductory group sessions, community group discussions and in-depth interviews (Table 1), with the intention that the different methods complemented each other. Fieldwork was conducted at Site A first, after which the field team moved to Site B.

**Table 1:** Description of study areas and characteristics of study participants

| Method of data collection | Data collected at Site A | Data collected at Site B |
| --- | --- | --- |
| **Transect spiral walk** | **Approximate population size:** 700 people<br>**Main economic activity:** Farming/trading<br>**Health facilities:** No health facility. Healthcare services sought from neighbouring villages.<br>**Healthcare providers:** Retired midwife, VHTs, Traditional Healer | **Approximate population size:** 2,000 people<br>**Main economic activity:** Trading<br>**Health facilities:** a medical centre, pharmacies/drug stores<br>**Healthcare providers:** Clinical Officers, Nurses, VHTs, Traditional Healers, Herbalists |
| **Observations** | Comments recorded from five individuals including traditional healers. | Comments recorded from four individuals, including healthcare workers and herbal medicine seller. |
| **Introductory group session** | Discussion with community leaders (6 males, 5 females), aged 35-77. | Discussion with community leaders (5 males, 5 females), aged 35-50. |
| **Community group discussions** | Discussion with young females (n=10, aged 18-30), older females (n=12, aged 35-70), young males (n=9, aged 20-30), older males (n=9, aged 40-70). | Discussion with young females (n=10, aged 20-35), older females (n=11, aged 35-70), young males (n=11, aged 18-30), older males (n=10, aged 35-55). |
| **In-depth individual interviews** | 11 individuals (5 males; 6 females), aged 20-50 | 8 individuals (3 males; 5 females), aged 30-50 |
| **Total number of participants** | **67** | **64** |

### Data collection

Data collection was done by two social science research assistants who have an understanding of these communities and are fluent in Luganda, the local language commonly used within the study setting. The research assistants were first trained on all aspects of the study and the use of a rapid qualitative assessment approach (14). Before visits to the study communities, the research teams met with local leaders and explained the purpose of the study and sought permission to enter these communities. All the data were recorded using field notes that were expanded immediately after data collection.

i. **Transect spiral walk:** the aim was to observe, discuss and find key features in the community that would inform our other methodologies such as identifying settlement patterns, health centres, churches, mosques, traditional healers, trading centres, bars and any other places where people within communities concentrate. The transect walk was performed to understand the daily activities of people within the community in order to plan for the other assessment methods effectively. Two members of the research team first visited the local leader of each study site and explained the reasons of the activity within this community. With the help of the local leader, the team identified the centre of each village that was used as a starting point. The team then split into two with each person taking a different direction. They moved in concentric circles until the whole village was covered. During the walk, observations were made in addition to talking to a few people.
ii. **Observations:** informed by the transect walk, we visited particular places of interest to make observations. The team engaged with members of the community and healthcare providers (including traditional healers and village health teams (VHTs), with the aim of better understanding the characteristics of individuals seeking health care, how and where they are treated, common illnesses, familiarity with HBV including local terms to describe it, its signs and symptoms and any personal experience of individuals with HBV infection.
iii. **Introductory group sessions:** we met with ten influential people within each community including community leaders and local religious leaders, who had been identified based on information collected during the transect walks. The aim of these sessions was to understand perceptions and beliefs of diseases and illnesses in the community, and to explore understanding and experiences of HBV infection. The sessions were moderated by one interviewer, while the other was observing and taking notes.
iv. **Community group discussions:** we facilitated group discussions among adolescents and adults to find out about perceptions and beliefs of disease within the community, specifically exploring understanding and experiences of HBV infection. Four discussions were held at each study site with ten participants in each group. These participants were identified after the first three visits within the study communities.
v. **In-depth individual interviews:** We interviewed a selected group of individuals who were willing to give information about their experiences, to allow deeper exploration of relevant issues established from the previous stages.

### Data management and analysis

Researchers involved with data collection provided detailed description of their observations and interviews in accounts written up from the notes. Two other researchers reviewed the transcripts prior to analysis for to check the quality and accuracy, and thereafter independently coded the transcripts. Themes and sub-themes relating to perceptions, beliefs and experiences of HBV and the description, and nature of associated stigma, based on narrative content and research questions. Quotes used in the article are words of the interviewee used in the accounts.

### Ethical approvals

Approval was provided by The Science and Ethics Committee of the Uganda Virus Research Institute (ethics no GC/127/18/05/645) and UK Oxford Tropical Ethics Committee, OxTREC, (ethics no. 516-18). All participants provided written informed consent for interviews and discussion groups. The local leaders provided verbal consent for the transect walks within their communities while health care providers, including traditional healers, consented for observations.

### Data availability

Our study data are available on-line in the form of supplementary data, comprising a table of metadata and statements collected from 131 participants (Table S1), and a summary of key themes raised by the study (Table S2). These are available on-line at https://figshare.com/s/0cac69e7e69ee1f94e65 (this link will be converted to a permanent DOI on acceptance of the manuscript for publication).

## RESULTS

### Study participants

Between September and December 2018, we collected data from a total of 131 participants across Sites A and B. Table 1 summarises the description of study areas and socio-demographic characteristics of study participants.

### Knowledge and awareness of HBV infection

Awareness of HBV infection in these communities was varied. The commonest means of acquiring knowledge on HBV infection was through radio advertisements which encouraged members of the community to get tested and immunised (reported by 31 participants, age range 18-70 years). Others had personal experience of knowing someone perceived to be infected or diagnosed with HBV (reported by 30 individuals, age range 18 – 70 years). Among these respondents, 20 reported the death of individuals perceived to have HBV infection shortly after admission to a healthcare facility or as soon as they became symptomatic. One personal experience was reported by a teacher from site B: *“A neighbour who had gone to work at the lake shore came back when he was already infected. He was referred to a hospital in Kampala but died after two days”*.

There were some participants who reported not having heard of HBV infection and therefore felt that it may not be a common illness, including both males and females from both settings. There is no specific name for HBV infection; naming of a disease in this community commonly depends on the symptoms of the disease. The local phrases used to describe liver disease are *“Obulwadde bwekibumba” and “endwadde y’ekibumba”* which literally translate to ‘the liver is sick’.

### Insights into HBV as a cause of liver disease

As the symptoms and signs of HBV are not distinct from other causes of liver disease, there may be poor understanding of the role of an infectious agent. This is illustrated from a comment recorded from the community group discussions at site A, “We do not know about its mode of transmission, signs and causes so we cannot tell if the disease exists in the community”. Three participants specifically reported that they are not able to distinguish HBV from other illnesses.

More generally, some participants were able to list other causes of liver disease including excessive consumption of alcohol and eating fatty foods. Insect bites, exposure to pesticides/fertilisers and drinking dirty water were also suggested as causes of liver disease. For example, a man at Site B commented that: *“it could be caused through excessive drinking of waragi [local gin]*…. *myrrh, khat drugs, herbal medicine, and fertilisers/pesticides can affect and make the liver sick”*, while a woman of a similar age in Site A suggested that it was caused by *“walking through dirty swamp water which contains certain germs that cause liver disease”*. Some of the symptoms that participants reported in association with HBV (or more generally with liver disease) included abdominal or whole body swelling, jaundice/yellow eyes, weight loss, and hair loss. Three participants mentioned that HBV was asymptomatic. Thirteen participants, mainly from Site A, were not able to suggest any signs or symptoms associated with HBV. Eight participants (aged 18-60 years) reported a similarity in signs and symptoms between HIV and HBV.

### Perceptions of mode of transmission

A number of participants, predominantly from Site B, reported that they did not know how HBV is transmitted. Three people believed that transmission routes included sharing of public toilets/bathrooms, or sharing of utensils and clothing. A male at site B stated: *“When he shares any equipment - say clothing, cups and forks with a person who looked malnourished - he would feel as if he was to contract the infection from him/her, not good to share anything with the patient”*.

Of the eight healthcare professionals who took part in this study, only five knew that HBV is sexually transmitted and can also be transmitted through other exposures to infected body fluids. Five participants from Site A reported that anyone was at risk of contracting HBV. People who consumed alcohol or those who engage in risky sexual behaviours were considered to be at risk of contracting HBV infection. One participant interpreted the high-profile campaigns around HBV immunisation for adults as suggesting that only adults were susceptible to infection.

### Knowledge of, and access to, HBV treatment

Community members suggested possible management interventions, including modified diet and herbal medicine. Twelve participants, mostly from Site A (age range 18-70 years), did not know any treatment for HBV. Three young men from Site B reported that HBV has no cure. Others correctly recognised that antiretroviral drugs can be used in managing HBV infection, and that those infected are put-on long-term treatment, but some of the comments suggested that there is pessimism about outcomes, with participants suggesting poor chances of survival.

### Potential challenges facing HBV management

The asymptomatic nature of HBV can be a barrier to seeking healthcare services; some community members only seek treatment when the symptoms are noticeable and have worsened, and it may be common to stop taking medication in the absence of symptoms. Poverty, preference for traditional/herbal medicine and lack of awareness about a disease, can also contribute to delays in seeking healthcare and be barriers to seeking preventive, diagnostic and treatment services among members of the community. A description of a child unwell as a result of HBV infection was provided by a male participant at Site A: *“When the boy fell sick, he was taken to the health centre for treatment. He was not admitted but they gave him some tablets which he didn’t understand. He took the tablets and after a week the stomach normalized. About 3 weeks later, the stomach resumed swelling. He was taken to herbal clinic and was given 5 litres of herbal medicine and he was told to go back after one week. When the herbal was finished, he went back and still he was given the same drugs. By that time his eyes had a yellowish colour, his urine was also yellow”*.

Lack of diagnostic facilities, and limited access to healthcare professionals and medications contribute to the challenges facing HBV prevention, diagnosis and therapy in this community. A female healthcare worker at site A explained: *“She can’t even give that patient any drug, only refers him or her to Masaka hospital. She said that they don’t have the machines that can diagnose it, so they have to refer the patient*.*”* Among healthcare workers, poor knowledge can impede diagnosis and treatment for HBV infection. Three healthcare professionals reported not having received a complete course of HBV vaccination themselves and are therefore at increased risk of becoming infected.

### Community perceptions and experience of HBV

We sought to understand the perception of community members about the occurrence and persistence of HBV infection in their community. Eleven participants, mostly from Site B reported that HBV was a new infection in their community because they had become aware of it only recently. For ten people who commented that HBV was a long-standing problem, their knowledge came from a community member known to them whom they believed had died of the infection. Others claimed that HBV was “an old sickness” which had been in the communities for a long time.

### Emotional responses to HBV infection

Communities tend to fear diseases based on symptoms and outcomes associated with the disease. In particular, a disease can be feared because it is incurable and may be seen as leading inevitably to death(4). Stigma may stem from this fear, as well as from changes in body appearance that can limit interactions with other community members. Eight participants, primarily from Site B, reported that they would be scared or more cautious to take care of someone with an infectious disease. Ten participants, mostly men from Site B (age 18–50 years), reported a feeling of shame associated with sexually transmitted diseases. However, eleven participants, reported no fear or shame towards people infected with HBV, partly because it is a disease that is not well known within the community.

## DISCUSSION

With the recent increased attention regarding the urgent need to upscale prevention, diagnosis and treatment of HBV infection globally, understanding the social impact of HBV infection in communities is important as this influences acceptability and impact of interventions. To date, there is very limited research on behaviour and beliefs surrounding HBV infection in Africa.

Familiarity with HBV infection in the communities we investigated can be explained, in part, by the recent efforts by the Ministry of Health in Uganda to combat transmission of HBV by promoting vaccination among adolescents and young adults (15), particularly using radio messages to reach a wide audience. As a result, the syndrome of HBV infection is now recognised by many adults in this setting. However, even among healthcare workers, there is still a lack of in-depth understanding, in keeping with similar studies in Ghana and Zambia (4,6–8). The importance of education is highlighted by a study of HBV infection undertaken in Turkey(16).

Naming of a disease can be a reflection of how much it is recognised and understood (17,18). In our study population, local names used to describe diseases are based on the symptoms the sufferer bears. In part, this practice is encouraged by the WHO which states that a name of a disease should be based on the ‘symptoms, disease manifestation, severity and, if known, the pathogen causing the disease’ (19). Naming HBV infection poses a challenge, since there is no specific symptom that distinguishes it from other liver diseases, and in many instances, it remains asymptomatic until the end-stages of infection. Although we identified local terms that broadly describe liver disease, gaps remain in understanding of HBV infection, particularly with regard to the infectious agent (19).

### Stigma

As a result of limited understanding and experience of HBV in our study population, stigma did not emerge as a strong theme. However, fear of symptoms and outcomes of a disease, as well as shame associated with mode of transmission of infection, can determine how those infected might be handled within their communities (20,21), and can lead on to stigma. Although sex is not the main route of transmission of HBV infection, this association could nevertheless lead to stigma and discrimination (22). Although HBV is asymptomatic in most cases, acute infection or advanced stages of the disease could result in visible manifestations, leading to isolation and lack of support from community members. Careful dissemination of accurate information on HBV is required to prevent possible stigma and misperceptions of personal risk.

Studies from Asia have reported stigma and limited knowledge among health practitioners as obstacles in the uptake of preventive, diagnostic and treatment services among HBV patients (4,23). Delayed health seeking behaviour has been widely reported in tuberculosis and is a result of several factors such as lack of symptoms, low patient knowledge, practising self-medication, and the use of traditional healing methods (24–26). In our study, the asymptomatic nature of HBV, poverty and lack of knowledge (including among healthcare workers) were challenges in accessing timely and appropriate healthcare (27). While there are efforts to improve knowledge on HBV among healthcare professionals and community members in Uganda, studies are needed to evaluate the evidence of effectiveness of these interventions, and to evaluate availability, accessibility and integration of HBV services.

### Strength and limitations of this study

This is the first study to explore beliefs, understanding and stigma around HBV infection in Uganda. A strength of the study was that we sampled participants from both urban and rural settings, sampling men and women and including a diverse range of ages which afforded a comparison for HBV description, experiences and understanding. This approach provides some valuable reflections and insights into local understanding and beliefs about HBV infection, but represents only a small number of adults, who self-selected to participate.

## Conclusions

Engaging local communities, understanding barriers, and providing appropriate and accessible information will be a crucial undertaking if diagnosis, treatment and preventive strategies are to be rolled out to support progress towards the 2030 sustainable development goal targets for elimination of HBV infection as a public health threat. Although radio campaigns have raised some local awareness of HBV in the communities we studied in Uganda, there remain significant gaps in knowledge and understanding, and a risk of stigma associated with fear, especially in the context of physical signs of liver disease. Educating traditional healers and healthcare workers could have a positive impact on disseminating information. Further studies are needed to investigate the societal responses to HBV infection in other African populations, and to establish the best ways to engage with local populations to promote access to interventions.

## Data Availability

Our study data are available on-line in the form of supplementary data, comprising a table of metadata and statements collected from 131 participants (Table S1), and a summary of key themes raised by the study (Table S2).

https://figshare.com/s/0cac69e7e69ee1f94e65

## SUPPLEMENTARY DATA

Supplementary data are available on line at Figshare: (https://figshare.com/s/0cac69e7e69ee1f94e65).

This will be converted to a permanent DOI on acceptance of the paper for publication.

**S1 Table: Data extraction summarising responses to discussion about knowledge, experience and beliefs about Hepatitis B virus infection from 131 study participants in Uganda**. Table summarises feedback from individuals and group discussions.

**S2 Tables: Common themes obtained from analysing responses obtained from study participants participating in discussion about knowledge, experience and beliefs about Hepatitis B virus infection in Uganda**. These represent responses from all individuals represented in the study. Note there is not a defined denominator, as there were differing approaches to approach discussion with study participants.

## ABBREVIATIONS

AIDS: Acquired Immunodeficiency Syndrome
GPC: general population cohort, Uganda
HBV: hepatitis B virus
HIV: human immunodeficiency virus
sSA: sub Saharan Africa
UK MRC: United Kingdom Medical Research Council
WHO: World Health Organisation

## ACKNOWLEDGEMENTS

Nil

## REFERENCES

1. World Health Organisation. Global hepatitis report, 2017. WHO. 2017;

2. World Health Organisation. Global health sector strategy on viral hepatitis 2016-2021.

3. Cooke GS, Andrieux-Meyer I, Applegate TL, Atun R, Burry JR, Cheinquer H, et al. Accelerating the elimination of viral hepatitis: a Lancet Gastroenterology & Hepatology Commission. Lancet Gastroenterol Hepatol. 2019;4(2):135–84.

4. Mokaya J, McNaughton AL, Burbridge L, Maponga T, O’Hara G, Andersson M, et al. A blind spot? Confronting the stigma of hepatitis B virus (HBV) infection - A systematic review. Wellcome Open Res. 2018;3:29.

5. O’Hara GA, McNaughton AL, Maponga T, Jooste P, Ocama P, Chilengi R, et al. Hepatitis B virus infection as a neglected tropical disease. PLoS Negl Trop Dis. 2017;11(10):e0005842.

6. Franklin S, Mouliom A, Sinkala E, Kanunga A, Helova A, Dionne-Odom J, et al. Hepatitis B virus contact disclosure and testing in Lusaka, Zambia: a mixed-methods study. BMJ Open. 2018;8(9):e022522.

7. Mkandawire P, Richmond C, Dixon J, Luginaah IN, Tobias J. Hepatitis B in Ghana’s upper west region: a hidden epidemic in need of national policy attention. Health Place. 2013 Sep;23:89–96.

8. Adjei CA, Naab F, Donkor ES. Beyond the diagnosis: a qualitative exploration of the experiences of persons with hepatitis B in the Accra Metropolis, Ghana. BMJ Open. 2017 Nov;7(11):e017665.

9. Ochola E, Ocama P, Orach CG, Nankinga ZK, Kalyango JN, McFarland W, et al. High burden of hepatitis B infection in Northern Uganda: results of a population-based survey. BMC Public Health. 2013;13(1):727.

10. Bayo P, Ochola E, Oleo C, Mwaka AD. High prevalence of hepatitis B virus infection among pregnant women attending antenatal care: a cross-sectional study in two hospitals in northern Uganda. BMJ Open. 2014;4(11):e005889.

11. Bwogi J, Braka F, Makumbi I, Mishra V, Bakamutumaho B, Nanyunja M, et al. Hepatitis B infection is highly endemic in Uganda: findings from a national serosurvey. Afr Health Sci. 2009;9(2):98–108.

12. Asiki G, Murphy G, Nakiyingi-Miiro J, Seeley J, Nsubuga RN, Karabarinde A, et al. The general population cohort in rural south-western Uganda: a platform for communicable and non-communicable disease studies. Int J Epidemiol. 2013;42(1):129–41.

13. Asiki G, Murphy GA V, Baisley K, Nsubuga RN, Karabarinde A, Newton R, et al. Prevalence of dyslipidaemia and associated risk factors in a rural population in South-Western Uganda: a community based survey. PLoS One. 2015;10(5):e0126166.

14. Bond V, Ngwenya F, Murray E, Ngwenya N, Viljoen L, Gumede D, et al. Value and Limitations of Broad Brush Surveys Used in Community-Randomized Trials in Southern Africa. Qual Health Res. 2019;29(5):700–18.

15. World Hepatitis Day 2018: Press statement on the progress of implementation of Hepatitis B vaccination program in Uganda https://reliefweb.int/report/uganda/world-hepatitis-day-2018-press-statement-progress-implementation-hepatitis-b

16. Tosun S, Aygün O, Özdemir HÖ, Korkmaz E, Özdemir D. The impact of economic and social factors on the prevalence of hepatitis B in Turkey. BMC Public Health. 2018;18(1):649.

17. Scully JL. What is a disease? EMBO Rep. 2004;5(7):650–3.

18. Shim K. Expanding the Definition of Infectious Disease. J Glob Health. 2015

19. World Health Organisation. WHO issues best practices for naming new human infectious diseases.

20. Mahajan AP, Sayles JN, Patel VA, Remien RH, Sawires SR, Ortiz DJ, et al. Stigma in the HIV/AIDS epidemic: a review of the literature and recommendations for the way forward. AIDS. 2008;22 Suppl 2(Suppl 2):S67–79.

21. Halli SS, Khan CGH, Moses S, Blanchard J, Washington R, Shah I, et al. Family and community level stigma and discrimination among women living with HIV/AIDS in a high HIV prevalence district of India. J HIV AIDS Soc Serv. 2016;1–16.

22. Blanas DA, Nichols K, Bekele M, Shankar H, Bekele S, Jandorf L, et al. Adapting the Andersen model to a francophone West African immigrant population: hepatitis B screening and linkage to care in New York City. J Community Health. 2015 Feb;40(1):175–84.

23. Lee H, Fawcett J, Kim D, Yang JH. Correlates of Hepatitis B Virus-related Stigmatization Experienced by Asians: A Scoping Review of Literature. Asia-Pacific J Oncol Nurs. 2016;3(4):324.

24. Mbuthia GW, Olungah CO, Ondicho TG. Health-seeking pathway and factors leading to delays in tuberculosis diagnosis in West Pokot County, Kenya: A grounded theory study. PLoS One. 2018;13(11):e0207995.

25. Chimbatata NBW, Zhou C-M, Chimbatata CM, Xu B. Post-2015, why delay to seek healthcare? Perceptions and field experiences from TB healthcare providers in northern Malawi: a qualitative study. Infect Dis Poverty. 2017;6(1):60.

26. Seid A, Metaferia Y. Factors associated with treatment delay among newly diagnosed tuberculosis patients in Dessie city and surroundings, Northern Central Ethiopia: a cross-sectional study. BMC Public Health. 2018;18(1):931.

27. Lemoine M, Thursz MR. Battlefield against hepatitis B infection and HCC in Africa. Hepatology. 2017; 66.

